# Prevalence and impact of foot peripheral neuropathy in Sytemic Sclerosis (SSc): results from a single centre cross-sectional study

**DOI:** 10.1101/2024.04.12.24305730

**Authors:** Begonya Alcacer-Pitarch, Marco Di Battista, Anthony C. Redmond, Anne-Maree Keenan, Stefano Di Donato, Maya H. Buch, Francesco Del Galdo

## Abstract

**Introduction:** Peripheral Sensory Neuropathy (PSN) is an under-recognized feature in systemic sclerosis (SSc). Moreover, SSc foot involvement is frequent but poorly investigated. We aimed to provide a detailed characterization of foot peripheral neuropathy in a large cohort of SSc patients, describing its associations with disease-specific features, physical disability and Quality of Life (QoL).

**Methods:** SSc patients and healthy controls (HC) comparable for age and gender, were enrolled in a cross-sectional observational case-control study. All subjects underwent a detailed quantitative sensory testing (QST) of feet evaluating touch, vibratory, thermal, and pain sensitivity; ultimately investigating the presence of large and small fiber neuropathy. Neuroptahtic symptoms were captured through a numerical rating scale assessing the presence of paraesthesia, numbness, burning, and stabbing pain. While the Manchester Foot Pain and Disability Index (MFPDI), SSc Health Assessment Questionnaire Disability Index (HAQ-DI), and the Systemic Sclerosis Quality of Life (SScQoL) needs-based questionnaire were used to capture the impact of the PSN on foot disability and QoL.

**Results:** 109 SSc patients (88.1% female, median age 59.0 years) and 51 HC were enrolled. SSc patients presented with a significant median reduction of areas with preserved tactile sensitivity (14 IQR 4; p<0.001), and a delayed vibration perception threshold (1.7 µm IQR 3.0; p=0.01). Regarding thermoreceptor impairment, they presented with signifiantlly higer cold and warm thresholds (27.0 °C, IQR 3.0; vs 28.2 °C, p<0.001; 38.4 °C IQR 4.6, p=0.003 respectivelly), greater warm-cold threshold range (11.2 °C, IQR 6.9, p<0.001), and higher heat-induced pain threshold (44.8 °C, IQR 3.5; p<0.001),. At group level, 85.3% patients showed PSN on the feet, with 80% having small fibre involvement and 57% having large fiber neuropathy; while the coexistence of the two was present in 51.4% of the cases. Leaving only 14% without neuropathy. From those patients with PSN, 80.6% reported at least one neuropathic symptom, while 18% were asymptomatic. PSN was associated with age, smoking, foot ulceration,disease duration and corticosteroids use. Patients with neuropathic symptoms reported worse physical function, worse foot disability, and poorer QoL.

**Conclusion:** Foot PSN presents as common and disabling manifestation in patients with SSc, involving both large and small fibers, often co-existing. Clinically, the presence of neuropathic symptoms might serve as an indicator of PSN, although it can have a subclinical presentation. Hence, PSN assessment should be included as part of the workup of the SSc patient .

## Introduction

Systemic sclerosis (SSc) is a chronic connective tissue disease whose pathogenesis is attributable to vasculopathy, autoimmune deregulation, and tissue fibrosis. It can potentially affect any organ, thus presenting a very heterogeneous clinical expression and determining a significant burden on the patient’s quality of life [1]. Growing attention is gathering around neurological involvement in SSc, which can be expressed both as central or peripheral neuropathy as well as autonomic dysfunction [2,3]. Among those neurological manifestations, peripheral sensory neuropathy (PSN) is an under-recognized feature that has been poorly characterized; this could be attributed at least in part to the variability in definitions used for its diagnosis. Nonetheless, several studies have shown that it is by no means an uncommon problem. According to the most recent systematic reviews the prevalence of PSN, accounting for highly variable definitions, ranges from 14.5% to 27.3% [2,4], mostly affecting cranial, truncal and upper extremities nerves. Although PSN of the lower extremities has been reported in several studies, it has only been investigated in small cohorts [5–8]. When sensory symptoms are present, they can vary from numbness, paraesthesia and allodynia to stabbing and burning pain,; however, a subclinical presentation of foot PSN has also been reported [5–7].

PSN in SSc patients can have several different aetiologies including ischemia, tissue fibrosis, nerve compression through calcinosis, traumatic injury, medication adverse effects, and comorbid conditions such as diabetes mellitus [4,9]. The pathophysiological mechanisms underlining PSN in SSc are not yet fully understood, but a significant reduction in the density of myelinated fibers was found in sural nerve biopsies from SSc patients with multiple mononeuropathy [10], whereas skin biopsies showed a loss of myelinated and unmyelinated sensory autonomic nerve fibres (A-delta and C-fibers) [5]. Even though classification by type of peripheral nerve fibre involved, pattern of distribution and time of onset may aid in diagnosis, such a broad array of possible causes can make peripheral neuropathy in SSc a clinical challenge for the physician to face.

It should also be added that, unlike hand problems which are well known, foot problems in SSc have been poorly investigated. However, the few published studies show that SSc foot involvement is frequent and often disabling. In fact, the vast majority of SSc patients were found to have various degrees of morbidity and disability in all aspects of the foot, from the presence of pre-ulcerative lesions to the absence of peripheral pulses, from calcinosis to radiological abnormalities, up to biomechanical compromise, and all these factors lead to a significant burden on the patient’s quality of life [11–13].

The aim of this study was to provide a detailed characterization of foot peripheral neuropathy in a large cohort of patients with SSc, exploring any association with disease-specific characteristics, and assessing the impact of foot neuropathy on foot disability and quality of life.

## Methods

Adult consecutive patients affected by SSc according to 2013 EULAR/ACR classification criteria [14] and attending a routine visit at the Scleroderma clinic of Leeds Teaching Hospital NHS Trust were enrolled for this cross-sectional observational study along with a group of healthy controls (HC) comparable for age and gender. HC recruitment was undertaken through SSc subjects, using a technique referred to as “bring a friend” where patients were asked to nominate a healthy friend/relative of the same gender and age (±2 years) who was willing to participate. This recruitment strategy has been described previously in the literature as an effective method of matching for socio-economic, ethnicity and other demographic factors. Ethical approval was obtained from the local ethics committee (Leeds Research Ethics Committee, ref 10/H1306/14) and all participants provided written informed consent to participate in accordance with the Declaration of Helsinki.

Demographic and SSc-specific variables and comorbidities were collected for each patient, including: disease duration, limited (lcSSc) and diffuse cutaneous (dcSSc) subset according to LeRoy [15] and modified Rodnan skin score (mRSS), autoantibody positivity distinguishing between anti-centromere and anti-topoisomerase I autoantibodies, history of ulcers specifying whether located on the foot or elsewhere in the body, current medications distinguishing between immunosuppressants, corticosteroids and vasoactive drugs. The presence of ongoing ulcers on the feet, diabetes mellitus and the history of orthopaedic or vascular surgery of the lower extremities in the previous 12 months were considered exclusion criteria. All enrolled subjects underwent a detailed quantitative sensory testing (QST) of feet. Patients were also questioned about the presence of neuropathic symptoms such as paraesthesia, numbness, burning or stabbing pain, according to a numerical rating scale (NRS) from 0 (no symptoms) to 10 (most severe symptoms). Finally, patient-reported outcomes (PROMs) were administered to assess physical disability, and patient’s quality of life.

### Quantitative sensory examination

The somatic sensory nerve fibres of both feet were examined for sensations of touch, vibration, temperature and pain with the different QST tools (*Somedic SenseLab, Sösdala, Sweden*). Specifically, QST investigated large diameter myelinated fibers (A-alpha and A-beta) which carry touch and vibration sense along with motor function, as well as small diameter myelinated (A-delta) and unmyelinated (C) fibers, which carry temperature and pain sensations along with autonomic function [9,16]. Patients were evaluated in a quiet and temperature-controlled room at 23 ±1.3 °C (relative humidity 36 ± 9%). During the assessments, patients were asked to close their eyes to avoid any possible visual influence.

*Mechanical threshold* for light touch was assessed using a calibrated nylon von Frey monofilament with a diameter of 0.26 mm, thus exerting a pressure of 7.3 g/mm^2^, on eight different sites for each foot (five fingertips, dorsum, heel and ball). The monofilament was perpendicularly applied three times for each site with a slight pressure that determines its arching. Abnormal light touch testing was defined as one or more sites failing to detect two out of three applications.

*Vibration threshold* was assessed with a 100 Hz vibrameter using the method of limits. It was determined by the detection of onset of the vibratory stimulus on a bony prominence, the dorsal area of the first metatarsal shaft. The amplitude of the stimulus was slowly increased (1 µm/s, from 0 to 399.9 µm) until the patient reported feeling it. The vibration perception threshold was measured three times for each foot with a 10 s rest between measurements, the mean value was then calculated. *Thermal threshold* and *heat-pain thresholds* were measured objectively using the Modular Sensory Analyzer Thermal Stimulator (*Somedic SenseLab, Sösdala, Sweden*). This computer-controlled device generates and documents response to highly repeatable thermal stimuli, such as warmth, cold, and heat-induced pain. These stimuli are transmitted through a thermode (25 x 50 mm) which can be either cooled or warmed, placed dorsally over the metatarsophalangeal joints of both feet. The method of limits was again used to detect the thermal threshold and thermal-pain threshold. First, the perception threshold test (setting “mix stimulus”) was used to detect thermal sensitivity thresholds. Starting from a baseline temperature of 32°C, five cold stimuli followed by five warm stimuli were administered at intervals of five seconds each. The stimulation rate was 1°C/s, whereas the return to baseline rate was 3°C/s. Patients were asked to press a switch that reversed the current at the precise moment they felt a sensation of cold or warm. The mean value of both thresholds was then calculated, and a warm-cold threshold range was finally determined. After that, for heat-pain threshold a single heat stimulation (reaching a maximum of 50°C) was administered, and the patients had to press a switch to stop it at the precise moment they felt a sensation of pain.

The pathological impairment of each type of sensitivity assessed with QST was defined according to the different cut-offs reported in the literature; where there were no cut-offs from healthy subjects, those derived from diabetic neuropathy were used [17–19]. Patients were then considered as having large fiber neuropathy if at least one of the tactile and vibratory sensitivities was found pathological. Small fiber neuropathy was defined as the presence of impairment in at least one of thermal (both warm and cold) and pain sensitivities.

### Patient-reported outcomes

The following PROMs were administered to assess the impact of PSN on physical disability and quality of life:

-Scleroderma Health Assessment Questionnaire (SSc-HAQ): daily functional activity was assessed by twenty items grouped into eight domains with the addition of five SSc-specific visual analog scales. Each item was converted in a score from 0 to 3, with higher scores corresponding to worse disability, and a final mean score was then calculated [20].
-Manchester Foot Pain and Disability Index (MFPDI): a 19-item tool developed to specifically assess foot pain and disability. Each item can score 0, 1 or 2 and the final score is their sum, with higher scores corresponding to greater foot pain and disability [21]. Disabling foot pain reflecting impaired physical function was considered when at least one of the ten MFPDI function items were experienced on most/every day, i.e. with a score of 2 [22].
-Systemic Sclerosis Quality of Life Questionnaire (SScQoL): a 29-item tool that measures the disease impact on health and wellbeing, and that was developed using a needs-based quality of life model. Each item can score 0 or 1, the final score is their sum, with higher scores corresponding to worse health-related quality of life [23].

### Statistical analysis

Categorical data were described by absolute and relative frequency, continuous data were non-normally distributed and therefore summarised by median and interquartile range. Mann-Withney U-test (two-tailed) and Fisher test were conducted to compare continuous and categorical variables among groups, respectively. False-discovery rate correction was applied for multiple comparisons. Spearman’s correlation coefficient was calculated to assess the association between continuous variables. Multivariable regressions were computed to evaluate the influence of the different variables on the parameters of interest. Statistical significance was set at 0.05. All analyses were performed with R software (*R Core Team 2023*).

## Results

### Study population

One hundred and nine SSc patients (88.1% female, median age 59.0 years) were enrolled in the present study along with 51 HC comparable for median age and gender percentage. Epidemiological and SSc-specific characteristics of the cohort are reported in Table 1. Most patients had lcSSc and more than half presented anti-centromere positivity. The median disease duration was 8.0 years and only 8 patients had a history of foot ulcers.

**Table 1.** Epidemiological and disease-specific characteristics of the cohort.

|  | **SSc (n = 109)** | **HC (n = 51)** |
| --- | --- | --- |
| Age, years* | 59.0 (51-66) | 52.0 (43-63.5) |
| Female | 96 (88.1%) | 43 (84.3%) |
| BMI, Kg/m<sup>2</sup> * | 25.9 (21.8-30.3) | 24.9 (22.1-28.7) |
| Smoking habit | 55 (50.5%) | 21 (41.2%) |
| - Current smoker | 14 (12.8%) | 5 (9.8%) |
| Disease duration, years* | 8.0 (4.0-14.0) |  |
| Skin subset |  |  |
| - dcSSc | 24 (22%) |  |
| - lcSSc | 85 (78%) |  |
| mRSS* | 2.0 (0.0-6.0) |  |
| Autoantibody |  |  |
| - anti-centromere | 61 (56%) |  |
| - anti-topoisomerase I | 14 (12.8%) |  |
| - other | 34 (31.2%) |  |
| History of foot ulcers | 8 (7.3%) |  |
| History of ulcers elsewhere | 34 (31.2%) |  |
| Immunosuppressants | 24 (22%) |  |
| Corticosteroids | 23 (21.1%) |  |
| Vasoactive drugs | 98 (89.9%) |  |

Data are reported in median (IQR)\* and n (%).
No statistically significant differences between SSc and HC for every item reported.
dcSSc: diffuse cutaneous SSc; lcSSc: limited cutaneous SSc; mRSS: modified Rodnan skin score.

### Neuropathic symptoms

One or more neuropathic symptoms were reported by 89 (82%) SSc patients, with a significantly higher prevalence of complaints for numbness (55%), paraesthesia (52%), stabbing pain (37%), and burning pain (35%) than HC (p<0.001 for all). Accordingly, patients had higher NRS scores in each domain (Figure 1, P<0.001 for all). Univariate analysis of each symptom against clinical features showed that burning pain was significantly associated with history of foot ulcers (p=0.009), whereas stabbing pain showed a weak negative correlation with disease duration (p=0.001; ρ=-0.359) and age (p=0.03; ρ=-0.214).

**Figure 1.**
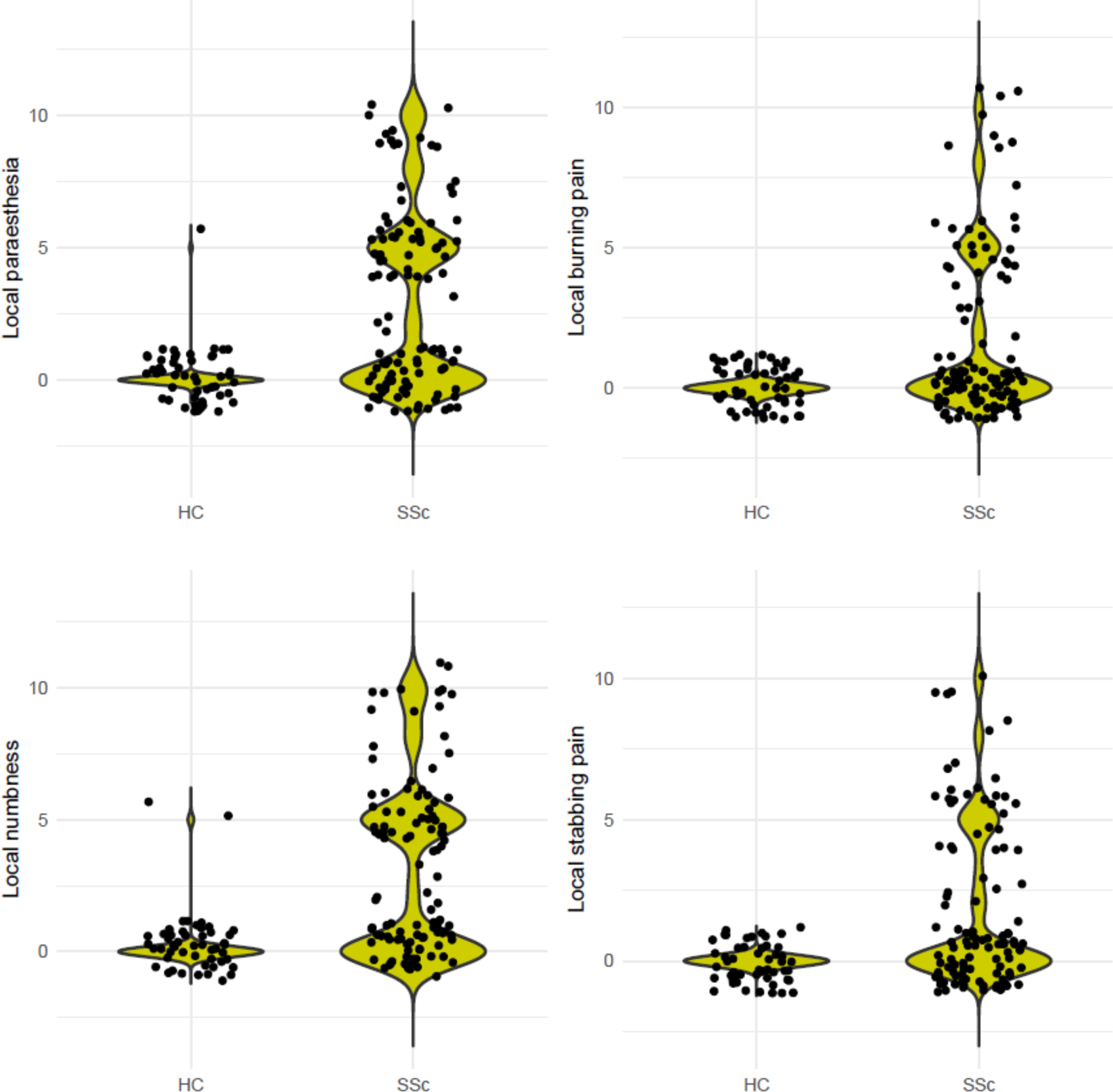
Differences in NRS scores for neuropathic symptoms between SSc and HC (p<0.0001 for all).

### Quantitative sensory examination

Consistent with patient reported PSN symptoms, sensory testing showed statistically significant differences between SSc and HC in all domains (Table 2). SSc cases presented with a significant median reduction of areas with preserved tactile sensitivity (14 IQR 4 vs 16 IQR 2; p<0.001) and a delayed vibration perception threshold (1.7 µm IQR 3.0 vs 1.1 µm IQR 1.3; p=0.01). There was a greater number of SSc patients with impaired tactile sensitivity (54% [n=59] vs 31% [n=16]; p=0.007), a similar trend was observed for pathological vibratory sensitivity with no statistical significance (12% [n=13] vs 6% [n=3]; p=0.2). When assessing thermal sensitivity, SSc patients showed a median cold threshold of 27.0 °C (IQR 3.0) which was significantly lower compared to 28.2 °C (IQR 2.9) from HC (p<0.001). Similarly, SSc cohort revealed a higher median warm threshold (38.4 °C IQR 4.6 vs 37.2 °C IQR 4; p=0.003). As a result, SSc patients presented a median warm-cold threshold range of 11.2 °C (IQR 6.9) which was significantly greater compared to 8.9 °C (IQR 5.9) from HC (p<0.001). The percentage of SSc patients with an impaired thermal sensitivity was meaningfully greater than HC (74% vs 47%; p<0.001).Lastlly, patients with SSc had a statistically significant reduction in pain perception when compared to HC (48% vs 27%; p=0.015). They presented with a median heat pain threshold of 44.8 °C (IQR 3.5), significantly higher compared to 43.1 °C (IQR 3.6) from HC (p<0.001), thus accounting for a greater reduction in pain perception in patients with SSc. .

**Table 2.** Quantitative sensory testing results for SSc patients and healthy controls.

|  | **SSc (n = 109)** | **HC (n = 51)** | **p-value** |
| --- | --- | --- | --- |
| Light touch mechanical threshold,<br>n. areas with preserved sensitivity* | 14.0 (12.0-16.0) | 16.0 (14.0-16.0) | **<0.001** |
| Tactile sensitivity impaired, n (%) | 59 (54) | 16 (31) | **0.007** |
| Vibratory threshold, $\mu\text{m}^*$ | 1.7 (1.0-4.0) | 1.1 (0.6-1.9) | **0.01** |
| Vibratory sensitivity impaired, n (%) | 13 (12) | 3 (5.9) | 0.2 |
| Large fiber neuropathy, n (%) | 62 (57) | 17 (33) | **0.006** |
| Warm thermal threshold, $^{\circ}\text{C}^*$ | 38.4 (36.7-41.3) | 37.2 (35.5-39.5) | **0.003** |
| Warm sensitivity impaired, n (%) | 89 (82) | 31 (61) | **0.005** |
| Cold thermal threshold, $^{\circ}\text{C}^*$ | 27.0 (25.3-28.3) | 28.2 (26.5-29.4) | **<0.001** |
| Cold sensitivity impaired, n (%) | 95 (87) | 41 (80) | 0.3 |
| Warm-cold threshold range, $^{\circ}\text{C}^*$ | 11.2 (9.0-15.9) | 8.9 (6.7-12.6) | **<0.001** |
| Thermal sensitivity impaired, n (%) | 81 (74) | 24 (47) | **<0.001** |
| Heat pain threshold, $^{\circ}\text{C}^*$ | 44.8 (43.1-46.6) | 43.1 (41.0-44.6) | **<0.001** |
| Pain sensitivity impaired, n (%) | 52 (48) | 14 (27) | **0.015** |
| Small fiber neuropathy, n (%) | 87 (80) | 28 (55) | **0.001** |

\*Data are reported in median (IQR)

On the basis of these results, the presence of PSN was found in 93 (85.3%) SSc patients (Figure 2A). In detail, 62 (57%) of them were classified as having large fiber neuropathy and 87 (80%) as having small fiber neuropathy (Figure 2B), in both cases with significantly higher percentages than HC (p=0.006 and p=0.001, respectively). More than half of SSc subjects had the co-presence of both large and small fiber neuropathy (Figure 2C).

**Figure 2:**
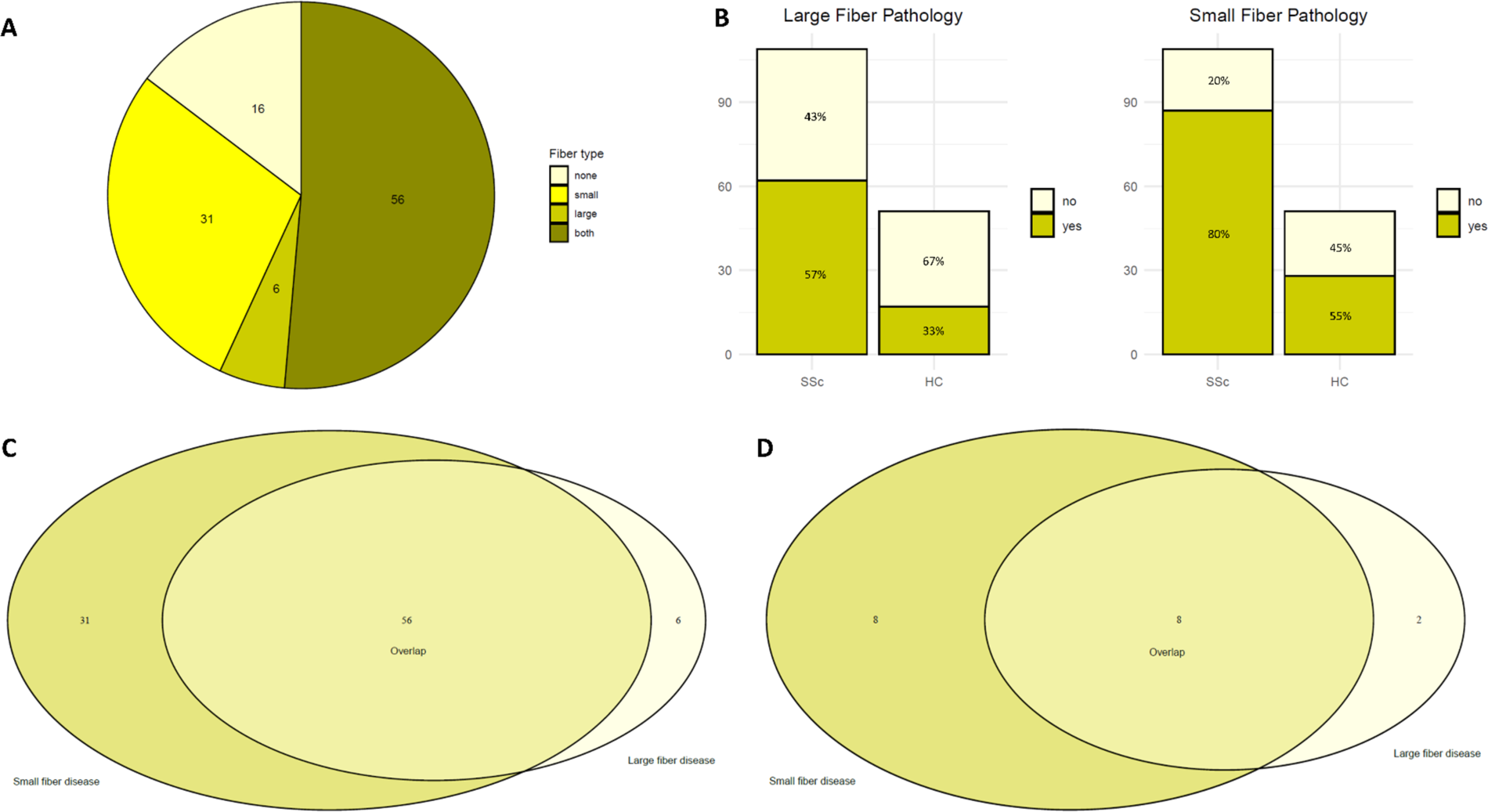
A) Distribution of fiber type neuropathy in SSc cohort (n=109), B) Prevalence of large and small fiber neuropathy across SSc and HC cohorts. C) Overlap of small and large fiber neuropathy across SSc patients presenting at least one type of nerve pathology (n=93). D) Overlap of small and large fiber neuropathy in asymptomatic SSc patients presenting at least one type of nerve pathology (n=18).

Out of 93 patients with PSN, 75 (80.6%) reported at least one neuropathic symptom. Interestingly, a subgroup of 18 (19.3%) SSc subjects who had PSN according to sensory testing did not report any neuropathic symptom. Ten (55.5%) of these patients had large fiber neuropathy, 16 (88.8%) had small fiber neuropathy, and 8 (44.4%) subjects presenting with both (Figure 2D).

### Association with disease characteristics

The relationship between sensory neuronal parameters and SSc disease characteristics was evaluated. We observed a weak negative correlation between age and the number of areas with preserved tactile sensitivity (p=0.003, ρ=-0.287). Vibration threshold was found significantly higher in patients with smoking habit (p=0.007) and history of foot ulcers (p=0.03), showing a moderate direct correlation with age (p<0.001; ρ=0.569). A multivariate analysis confirmed the association of all these variables with vibration sensitivity (B=2.16, p=0.006 for smoking habit; B=8.87, p<0.001 for history of foot ulcers; B=0.12, p<0.001 for age). Warm sensitivity threshold was significantly associated with smoking habit (p=0.01) and treatment with corticosteroids (p=0.009). Moreover, a direct weak correlation was found with age (p=0.003; ρ=0.279) and disease duration (p=0.04; ρ=0.219). After a multivariate analysis, only corticosteroids and age were confirmed significantly associated with warm sensitivity threshold (B=1.53, p=0.04 and B=0.06, p=0.01, respectively). On the other hand, the only correlation found for cold sensitivity threshold was with age (p=0.006; ρ=-0.261). As a result, warm-cold threshold range was significantly associated with smoking history (p=0.01), corticosteroids treatment (p=0.04) and weak correlated with age (p=0.001; ρ=0.298). In the multivariate analysis for warm-cold threshold range, only age was confirmed (b=0.13, p=0.006). Lastly, heat pain threshold was found in a direct weak correlation with age (p=0.007; ρ=0.253) and disease duration (p=0.03; ρ=0.210), both confirmed at a multivariate analysis (B=0.04, p=0.01 and B=0.05, p=0.04, respectively). No associations were found between SSc sensory neuronal results and gender, autoantibody positivity, skin subset, and treatment with immunosuppressants or vasoactive drugs.

When considering the presence of large and small fiber neuropathy in SSc cohort, patients affected were found to be significantly older (p<0.001 and p=0.004, respectively), especially when the two conditions co-existed (p<0.001). Small fiber neuropathy was more frequent in patients with lcSSc and ACA positivity (p=0.03 for both). No other relevant associations were found between the presence of large and/or small fiber neuropathy and epidemiological or disease-specific characteristics.

### Impact on physical function and quality of life

The relationship between PSN and physical function and quality of life was then evaluated by different PROMS. SSc-HAQ revealed a median score of 1.25 (IQR 0.54-1.76) . Subgroup analysis showed that SSc-HAQ was significantly higher in patients complaining of neuropathic symptoms such as paraesthesia (1.47 [IQR 0.95-1.89] vs 0.74 [IQR 0.46-1.38]; p=0.003), numbness (1.47 [IQR 0.88-1.90] vs 0.70 [IQR 0.45-1.38]; p<0.001) and stabbing pain (1.42 [IQR 0.75-1.86] vs 1.07 [IQR 0.45-1.73]; p=0.02). Similar results were obtained for MFPDI that yielded a median score of 20 (IQR 8-26) and was significantly higher in patients complaining of paraesthesia (22 [IQR 14-28] vs 18 [IQR 4-22]; p=0.01). Interestingly, according to MFPDI, 82 (75.2%) patients were affected by disabling foot pain (MFDPI > 4), and these patients were complaining more often of paraesthesia (p=0.007), numbness (p=0.006) and stabbing pain (p=0.04). SScQoL showed a median score of 16 (IQR 8-22) and was significantly higher in patients complaining of paraesthesia (21 [IQR 11-24] vs 12 [IQR 6-20]; p=0.009), numbness (21 [IQR 11-24] vs 11 [IQR 5-19]; p=0.003) and stabbing pain (21 [IQR 12-23] vs 12 [IQR 5-21]; p=0.008). SScQoL also presented a weak negative correlation with cold sensitivity threshold (p=0.008; ρ=-0.249). Apart from this, no other associations were found between PROMs and QST results or the presence of large/small fiber neuropathy.

## Discussion

Peripheral neuropathy and foot involvement are two under-recognized aspects of SSc, but both are actually more frequent than one might think [4,11]. Moreover, they are both able to independently determine disability in SSc [13,24]. The aim of our work was therefore to characterize PSN in SSc foot, then explore any association with disease-specific characteristics and assess its impact on physical disability and quality of life.

In our cohort, SSc patients presented a significant alteration of all the sensory parameters examined in comparison with HC. Schady *et al*. were among the first to look for peripheral neuropathy in SSc: they performed a complete QST in hands and feet of 29 patients but found convincing signs of neuropathic impairment only for tactile and thermal sensitivity [6]. More recently, Frech *et al*. showed in 20 SSc patients a significant foot vibratory alteration when compared to HC and a trend for tactile impairment [8], which instead was found significant in the hands of 15 SSc subjects [25]. To the best of our knowledge, this is the largest SSc cohort undergoing a complete neuropathic assessment for small fiber and large fiber neuropathy of the foot, and our results confirm the presence of PSN already suggested in smaller cohorts. Our results highlight that PSN in SSc often affects small fibres, thus corroborating the several works that have reported a pathological impairment in thermal and nociceptive sensitivity, as well as in autonomic dysfunction [3,7,16]. Moreover, the co-presence of large and small fibre neuropathy was even more frequent, thus strengthening the relevance and the potential severity of PSN in SSc. Large fiber neuropathy also has been previously reported in symptomatic and asymptomatic SSc patients [9]. When symptomatic, the most prevalent complain in our cohort was numbness and parasthesia, two symptoms commonly reported in clinic. It is also interesting to note that clinically asymptomatic patients were very often affected by large and/or small fiber neuropathy, so that subclinical PSN should be suspected even in SSc subjects not complaining of paraesthesia, numbness, stabbing or burning pain.

When evaluating associations with disease characteristics, age was the most impactful variable for almost all neurosensory parameters examined. These findings are in line with the well-recognized role of age in peripheral sensitivity [26]. While disease duration, smoking, history of foot ulceration and corticosterois use were signifiancatlly associated with one of the six sensory tests. We believe the significant association of smoking history and history of foot ulcers with vibratory sensitivity and the corelation with thermal sensitivity offers an interesting glimpse into the potential direct relationship between peripheral vasculopathy and neuropathy, re-proposing a pathogenetic model of interrelation already present in diabetes [27]. Furthermore, the association with corticosteroid treatment could be related to animal findings that prolonged exposure to glucocorticoids often leads to maladaptive neuronal and glial plasticity consisting of both structural and functional changes, particularly regarding mechanical allodynia and thermal hyperalgesia, associated with the development of neuropathic pain [28,29]. On the other hand, we observed an association between small fiber neuropathy and both lcSSc and anti-centromere positivity, not confirmed for large fiber neuropathy or their co-presence. In this context, there are contrasting results in the literature. In fact, there are studies that identify dcSSc and anti-centromere positivity as major risk factors for peripheral neuropathy [4,30], whereas others found a significant association with lcSSc and positivity for anti-topoisomerase I and anti-U1-RNP [9,31]. Although, there are also studies that have not highlighted a particular influence of cutaneous involvement or autoantibody profile [6,32]. In this regard, larger cohort, multicentre studies are needed to confirm association with specific diseae characteristics.

From the evaluation of different PROMs it clearly emerged that neuropathic symptoms as paraesthesia, numbness and stabbing pain are associated with worse physical disability and quality of life. These findings confirm, expand and enrich what was observed by Ivanova *et al* [24], whom observed that the severity of of neuropathic symptoms are associated wth a worse physical disability. On the other hand, the lack of significant associations between PROMs and QST results can be traced, rather than a mere lack of influence of peripheral neuropathy on quality of life, to the fact that PROMs are designed to reliably return the picture of subjective symptoms, while they may perform less well in capturing objectifiable instrumental alterations. However, it should be noted that MFPDI showed that three quarters of the SSc cohort is affected by disabling foot pain, and when compared to those without disaling foot pain the predominant neuropathyc symptoms where parasthesia, numbness and sabbing pain.

The main limitation of our work is the absence of nerve conduction studies to corroborate large fiber neuropathy, and skin biopsis to corroborate small fiber neuropathy and obtain a histologic evaluation. Nontheless, QST is recommended as a battery of tests to assess small fiber neuropathy [33]. The lack of use of the clinical total neuropathy score limits the comparison of our results with those of other similar studies that applied this assessment tool. Additionally, other underlaying diseases or causes that could contribute to the PSN, for example hematologic abnormaities, Vitamin B12, B1 and B6 deficiency were not assessed.

In conclusion, we showed that foot peripheral neuropathy is a common clinical entity and represents a concrete burden on physical disability and quality of life. However, routine screening for peripheral neuropathy is often not mentioned in best clinical practice guidelines [34]. Moreover, despite SSc patients have a relatively high prevalence of self-reported foot problems, their foot health care and information are usually inadequate [35]. These concerns are even more important when considering that SSc patients often present with a postural imbalance and a consequent increased risk of falls [36]. This work ultimately highlights the importance to screen all SSc patients for PSN, assessing the potential presence of both small and large fiber neuropathy.

## Conclusion

Foot PSN presents as common and disabling manifestation in patients with SSc, involving both large and small fibers, often co-existing. Clinically, the presence of neuropathic symptoms is associated with poorer physical function and QoL. Additionally, almost 1 in 5 patients with sensory testing proven PSN do not recognise their symptoms, supporting the role of PSN assessment as a valuable tool in routine SSc management.

## Data Availability

All data produced in the present study are available upon reasonable request to the authors

